# Investigator-initiated versus industry-sponsored trials – Visibility and relevance of randomized controlled trials in clinical practice guidelines (IMPACT)

**DOI:** 10.1101/2025.01.13.25320444

**Authors:** Manuel Hecht, Anette Blümle, Harald Binder, Martin Schumacher, Nadine Binder

## Abstract

**Background:** The goal of evidence-based medicine is to make clinical decisions based on the best available, relevant evidence. For this to be possible, studies such as randomized controlled trials (RCTs), which are widely considered to provide the best evidence of all forms of primary research, must be visible and have an impact on clinical practice guidelines. We further investigated the impact of publicly and commercially sponsored RCTs on clinical practice guidelines by measuring direct and indirect impactful citations and the time to guideline impact.

**Methods:** We considered the sample from the IMPACT study, where a total of 691 RCTs (120 German investigator-initiated trials (IITs), 200 international IITs, 171 German industry-sponsored trials (ISTs) and 200 international ISTs) was sampled from registries and followed prospectively. Their publications in the form of original articles and systematic reviews (SRs) citing these articles had already been identified, as well as clinical practice guidelines (CPGs) which cited either the articles or the SRs. The methods and results of this effort were already published. In this investigation we aimed to better characterize the impact of RCTs in CPGs. Therefore, we identified all citations of the original articles and SRs in the citing CPGs and classified them into impactful and non-impactful. This allowed us to calculate an estimate for the guideline impact of a trial. In addition, we estimated the time-to-guideline-impact, defined as the time to a direct and indirect impactful citation in a CPG. Direct means that the publication of a trial was cited in the main text of a CPG. Indirect means that the publication was cited and included in the findings of a SR and the SR was cited in the main text of a CPG. We also investigated to what extent pre-defined study characteristics influenced the guideline impact using multivariable negative binomial regression as well as the time-to-guideline impact using multivariable Cox proportional hazards regression.

**Results:** Overall, 22% of RCTs impacted a CPG. For international ISTs, only 15% of trials had an impact in CPGs. Overall, of the 405 associated guidelines, 331 were impacted. Larger trials were associated with more impactful main text citations in CPGs and earlier time-to-guideline impact, while international industry-sponsored trials were associated with smaller impact on CPGs and longer time-to-guideline impact. IITs funded by governmental bodies in Germany reached an impact on CPGs that is on par with German ISTs or international IITs and ISTs.

**Conclusion:** This study demonstrated that a considerable number of trials previously identified as being linked to CPGs have had impact in those CPGs (85%). International ISTs seem to have a lower impact on CPGs, and fewer of them influence CPGs at all.

## Introduction

Randomized controlled trials (RCTs) form the backbone of evidence-based medicine. They are the gold standard to detect causal relationships in medicine and to demonstrate the superiority of one treatment over the other [1]. However, they are also immensely resource intensive – ranging from the need of highly trained personnel, detailed planning, consent of and eventual hardships for the subjects as well as considerable amounts of money [2]. Given these facts, maximum visibility is desirable if not a moral necessity for the results of RCTs. Also due to the limited resources available, it is of great importance for public sponsors of RCTs to be able to target the most promising projects, e.g., in terms of considerable expected population-wide health benefit. Therefore, it is essential to assess the impact of RCT results in the medical literature. This impact was comprehensively described by the Becker Research Library Model [3] and according to Nury et al. [4] can essentially be broken down into the impact of the trial via publications, systematic reviews (SRs), or clinical practice guidelines (CPGs). Since it is highly challenging for clinicians to identify, read, and evaluate every publication of potentially relevant clinical studies in their field, the inclusion of RCT findings in SRs and CPGs is crucial for these findings to effectively influence patient care and treatment decisions. The time-to-impact of RCTs on the medical literature, especially CPGs, is also of primary concern. For example, there is a moral obligation for a drastically better treatment choice to become standard of care as soon as possible after careful examination of the evidence. The IMPACT study was designed and carried out with these objectives in mind [4, 5]. Specifically, an objective was to assess the visibility and impact of RCTs in the medical literature with respect to their sponsor type and primary study site. The study identified a total of 947 scientific articles resulting from 691 included RCTs, 2631 associated SRs, and 427 associated CPGs [5]. Medication studies and larger studies were more often published and cited compared to non-medication studies and smaller studies. The results of IITs were more frequently published as scientific articles, while the results of ISTs were more commonly found in study registries. German IITs were significantly inferior in terms of time from trial initiation to original publication of results compared to other cohorts. Additionally, smaller studies had a longer time to publication compared to larger studies [5]. A total of 26% of all trials were associated with CPGs (ranging from 16%-31% in between the cohorts). They were associated with CPGs mostly via 1st: result article only or 2nd: via SR and result article and this was stable over all cohorts (see Tab. 5 and Fig 7 from [5] respectively). The aim of this study was to further characterize the impact of the 691 RCTs in CPGs, especially to find a way to quantify the impact and the time it took for the trial results to gain impact in a CPG, hereinafter referred to as “time-to-guideline-impact”. To the best of our knowledge, there are no similar studies, which attempt to assess the impact on CPGs of RCTs in a prospective manner.

## Methods

### Study sample

We considered the sample from the IMPACT study, where - in 2018 - 691 multi-center RCTs were included from primary study registries with study start after 1st January 2005 and study completion before 31st December 2016, and specifically sampled according to four groups (i) German investigator-initiated trials (IITs) (n=120), (ii) international IITs (n=200), (iii) German industry-sponsored trials (ISTs) (n=171) and (iv) international ISTs (n=200). The 120 German IITs consisted of all RCTs sponsored by the two biggest independent public sponsors of trials in Germany: DFG (n=60) and BMBF (n=60). The RCTs were characterized by several measures such as the study size, number of primary outcomes, drug trial (yes/no), study phase (analogous to study phases in drug development) and medical domain. Subsequently publications of these trials were identified as results published in the trial registry or in the form of original articles published in scientific journals. Web of Science and NCBI PubMed were automatically searched for SRs citing the identified journal articles. Guideline databases (AWMF, TRIP and NICE) were searched automatically and manually for CPGs potentially citing either a trial registry number, an identified journal article, or an identified SR. These surveys took place from 2018 to 2019. All citations were validated. Further details of the study design have been published in a previous article [4].

### Guideline retrieval and publication date

For identifying the citations of publications and SRs from our cohorts, we acquired all CPGs in full text. We defined the publication date of a CPG as the earliest date when the guideline-text was publicly visible. If the guideline was a journal article, this was the earliest of either the electronic or the print publication date. If the guideline was published as a website or a PDF on a website, we set the publication date as the earliest date the guideline could be accessed in the web. In case of missing information on publication month or day, this was imputed via randomly drawing from the interval of possible values (eg. “1” up to “31” for days in January).

### Quantifying impact on CPGs as sum of citations

For each guideline, we searched the entire guideline-text for citations from our trial cohort. Figure 1 lists all possible paths from an RCT to a CPG in our data set: The CPGs could cite the trial registry number, the publication of a trial in form of a journal article or an associated SR. From here on onward we refer to the publications of the RCTs in form of an original article as *article*. In each step of citation, we differentiated between inclusion and exclusion. For citation of articles in SRs, the citations were checked manually in the IMPACT-study and were classified into *included*, *excluded* or *other*, the latter which corresponds to citations in the introduction or summary of a SR. Further details can be found in [4]. In this study, we postulated that the findings of an RCT can only be reflected in statements within a CPG if the RCT article was incorporated into the SR. Therefore, we defined only such citations as included, categorizing all other citations within SRs as excluded.

**Fig. 1:**
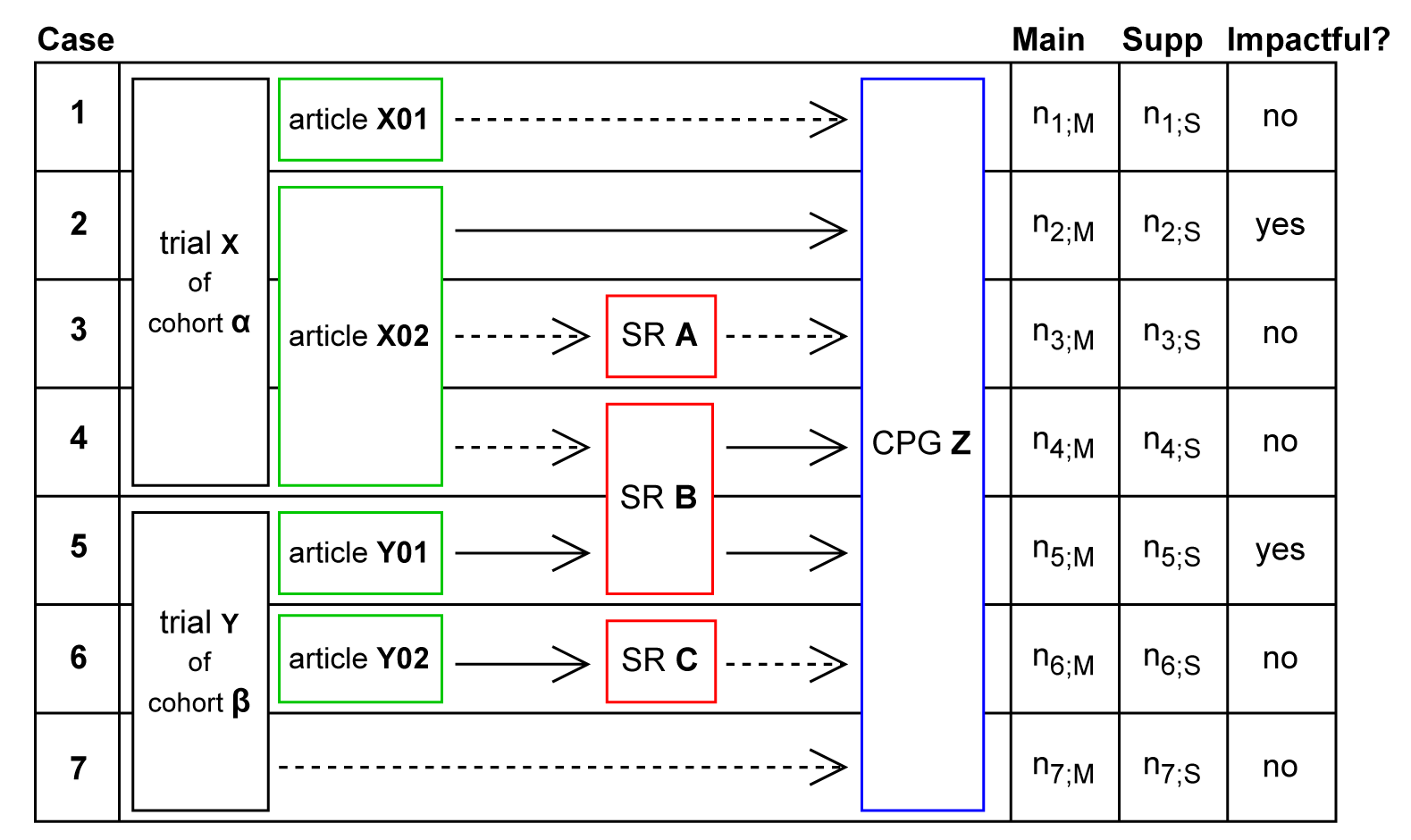
Pseudo-tabular data representation. Displayed are - from left to right: the presence or possible absence of entities like articles and SRs in between the possible connections between RCTs and CPGs in our data, the number of citations of the referenced entity in different parts of the CPG: Main Text (Main) or Appendix/Supplement (Supp) and whether a path was impactful. We displayed all possible paths from RCTs to CPGs (from top to bottom): Case 1: the CPG cited an article, but the article is excluded in the CPG (symbolized by dotted arrow), Case 2: the CPG cited an article and the article is included (symbolized by solid arrow), Case 3: the CPG cited but excluded or Case 4: included a SR and the SR cited an article but excluded it, Case 5: the CPG cited and included or Case 6. excluded an SR and the SR cites and included it, Case 7: the CPG cites the trial directly e.g., the trial registry number. In all these cases, the reference was not included in the CPG in our dataset, therefore we do not show the hypothetical case of a direct RCT citation with inclusion in CPG here. Inclusion in guideline was defined as having at least one citation in the meaningful sections of a guideline, thus ignoring the citations in the Appendix/Supplement (this only holds for cases 2, 4, and 5). We defined a path to CPG as impactful when in each step of citation the cited reference was included in the citing entity (SR or CPG).

For the citations in CPGs, we searched the full text for each mention of e.g. the citation number and classified these citations into citations in the main text or citations in the supplement (including the appendix). We defined a cited entity – we will call this a *reference* from here on onward – as included in a CPG, if it was cited in its main text. We defined a path from an RCT to a CPG as each unique observed connection through citations from an RCT to a CPG in our data set. Each unique path is illustrated by a single row in Figure 1. We defined a path as *impactful* if in each citing step the reference is included in the SR or CPG. There existed two possibilities for such paths:

1. In case the path from the RCT to the CPG did not involve an SR, it became *impactful* when the article was incorporated into the CPG.
2. In case the path from the RCT to the CPG included an SR in-between, it became *impactful* if the article was included in the SR and the SR was incorporated into the CPG.

In Figure 1, these cases correspond to rows 2 and 5 (solid arrows only). We define a path as *non-impactful*, when in at least one step from trial to CPG a citation was excluded (rows 1, 3, 4, 6, and 7 in Figure 1). We referred to a main text citation in a CPG that followed an impactful path as an *impactful guideline citation*. The *impact-on-CPGs*, i.e., the impact of an RCT on CPGs, was defined as the total number of its impactful guideline citations. Conversely, the number of distinct CPGs influenced by a trial, referred to as *impacted-CPGs per trial*, was defined as the count of unique CPGs with an impactful path connecting the trial to the CPG. The impact-on-CPGs as well as the impacted-CPGs were both 0 if there were no impactful paths for the RCT in question. We defined a binary status variable, guideline-impact (1 for impactful, 0 for non-impactful). Also we measured the *time-to-guideline impact* as the time from trial start to the earliest publication date of a CPG whilst only considering CPGs with an impactful path between CPG and the RCT in question. If an RCT had no impact-on-CPG, we treated the RCT to be right-censored on the 31st August 2019, corresponding to the last look-up-date for guidelines in the last search process for CPGs [5]. Apart from the guideline publication date and the citations we also measured the following guideline characteristics: the emitting society/societies, the guideline country or region, whether the guideline was a journal article itself, whether the guideline graded the evidence it cited, highlighted its recommendations, and whether it linked recommendations with the underlying evidence.

### Statistical analysis

We modeled the outcomes impact-on-CPGs and impacted-CPGs with multivariable negative binomial regression, including predefined relevant RCT characteristics. These models took the logarithm of the observed time per trial into consideration in the form of offset-terms. The observed time was defined as the time from study start to the end of the search period for CPGs (defined as the 31st August 2019). For investigating the time-to-guideline impact we provide Kaplan-Meier estimates for each cohort and multivariable Cox proportional hazards regression. In the sensitivity analysis we did not dichotomize baseline characteristics such as study. For reasons of readability, we will throughout this paper present values or estimates “X” that we wish to present for each cohort separately in the following manner (without providing the indices every time): *X*_(all trials)_ [X_(German IITs)_, *X*_(International IITs)_, *X*_(German ISTs)_, *X*_(International ISTs)_]. The alpha level was set to 5% in this study.

## Results

### Description of CPGs

For 95% of the 405 CPGs, one or several societies or institutions could be identified to be responsible for the guideline. In the other cases, only individuals could be identified. There was a wide variety in societies and authors, such that the most common societies (1st the American Heart Association (3%), 2nd. the AHA and American College of Cardiology (2%) and 3rd the Scottish Intercollegiate Guidelines Network (SIGN) (2%)) only emitted a small portion of the identified CPGs. Overall, 51% of CPGs were published in a scientific journal, whereas the remaining 49% were published elsewhere, e.g. on institutional websites or as conference abstract. The three most common journals publishing CPGs were Circulation (5%), European Heart Journal (3%) and the Journal of the American College of Cardiology (2%). The three most common countries or regions the CPGs derived from were the USA (24%), Germany (24%) and Europe (11%). Of all CPGs 73% systematically evaluated their underlying evidence, 85% clearly marked recommendations and 29% of all CPGs clearly linked their recommendations with the underlying evidence.

### Level of individual trials

At the level of individual trials, our data enable a very detailed visualization of the impact of a study on the literature in terms of transition plots. One such example illustrating the data complexity is given in Figure 2 (three further examples are provided in the supplementary material). Here one can see that the first published article is a methods article and is not included in the first SR it is cited in, but is included via a direct citation in the CPG citing this SR (corresponding to case 4 in Figure 1). In addition, the same article is included in a later SR. It is, however, the second published article and first result article of the trial to result in guideline-impact first (corresponding to case 2 in Figure 1). Also, this second article is referenced a lot more compared to the following four result articles. In this example, the trial tested spironolactone in patients with heart failure with preserved ejection fraction, and the first result article reported the effect on the primary outcomes: diastolic function and exercise capacity.

**Fig. 2:**
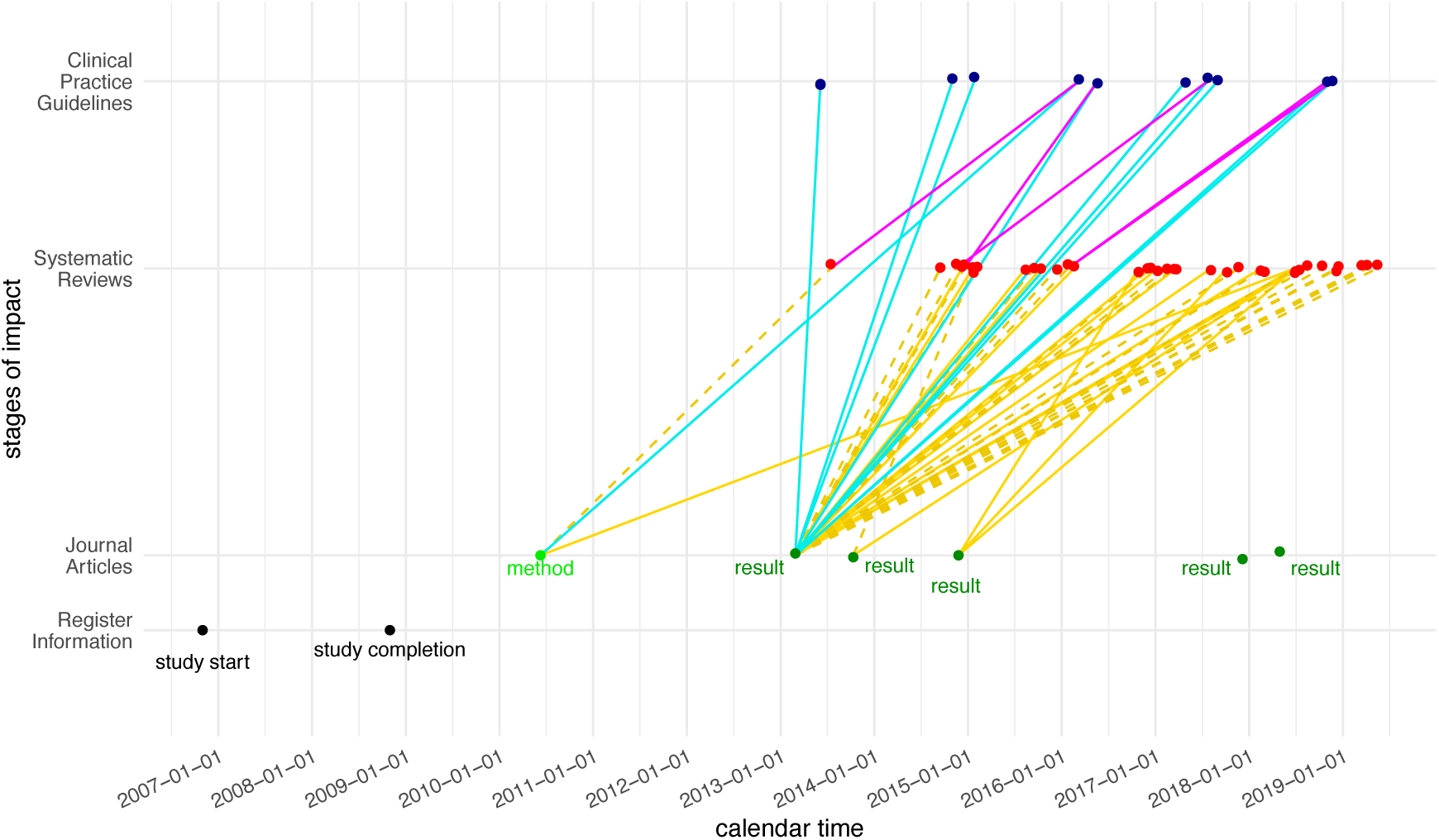
Stages of impact for one trial of the IMPACT study. From bottom to top, we displayed the information from the trial registry, then the publications as journal articles, the SRs citing these articles and the CPGs citing either the articles or the SRs. Dotted lines indicate exclusion in the citing entity, solid lines indicate inclusion. To be able to visually differentiate between CPGs citing SRs and articles these connections are color-coded with purple and blue respectively. Additionally, we displayed for the articles, whether they are a method article (only describing the methodology of a trial) or contained results.

The subsequent articles do not report the primary outcomes and could be described as further findings. This trial exhibits many commonly found patterns in this data set. For trials with associated SRs and CPGs, we found it to be typical that only a few of their articles were the major source for subsequent literature impact and for the first article to have been a method article and less referenced. The maximum number of articles for a single trial was 21. This was a trial examining the use of intra-aortic balloon pump (IABP) in cardiogenic shock due to myocardial infarction which published several sub-studies. The respective trial was associated with 60 SRs and 16 CPGs. It had impact on 15 of those CPGs, unsurprisingly so because the trial provided strong evidence against the usage of IABP in this indication. This was also the trial with the most associated and with the most impacted CPGs. In a dataset where only 22% of trials had any impact on CPGs, trials of this kind yielded a mean of 2.05 impact-on-CPGs per trial and 0.64 impacted CPGs per trial, with variances of 55.81 and 3.19, respectively. A histogram for the two outcomes *impact-on-CPGs* and *impacted-CPGs* is shown in Figure 3. Overall, 538 trials [89,149,130, 170] (77% [74%,74%,76%,85%]) did not gain any impact on CPGs. In total, we identified 2002 citations in CPGs, classified into 1893 citations in main text and 109 in the supplement.

**Fig. 3:**
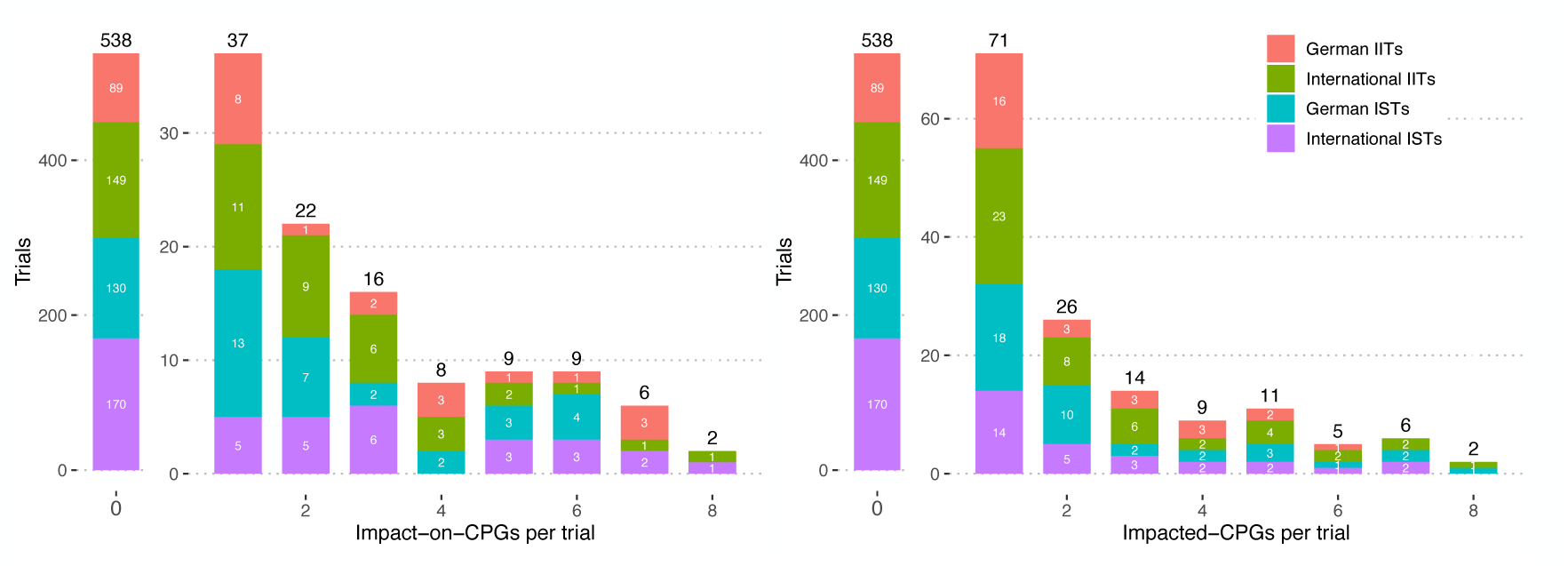
Histograms for the outcomes impact-on-CPGs (left) and impacted-CPGs per trial (right). Displayed are the absolute values, which should be interpreted relative to the cohort sizes 120, 200, 171, and 200, respectively.

### Lifecycle of trials

Figure 4 provides a detailed representation of the possible paths shown in Figure 1, along with concrete numerical data. In brackets the values are given for each of the four cohorts separately. About 68% (472) of all trials published their findings in a scientific article, 360 trials were then cited with their publications in SRs (946 distinct articles and 2628 distinct SRs were identified). Among the published trials, 238 were cited in a SR but ultimately excluded, while 309 were cited and included. One notable finding is that a large part of articles is cited in SRs, but only 44% percent are included in the citing SR. We note here that adding up excluded and included trials or articles provides only an upper bound on the number of distinct articles in this dataset that are associated with SRs. This is because a single trial may, through its articles, be included in one SR but excluded from another. The same reasoning also applies to how articles are associated with CPGs and how SRs are associated with CPGs. Of the 177 RCTs (26%) with a path to a CPG, only for 153 [31,51,41,30] (22% [26%,26%,24%,15%]) the path was impactful. The total number of impacted CPGs was 331 (82% of all associated CPGs).

**Fig. 4:**
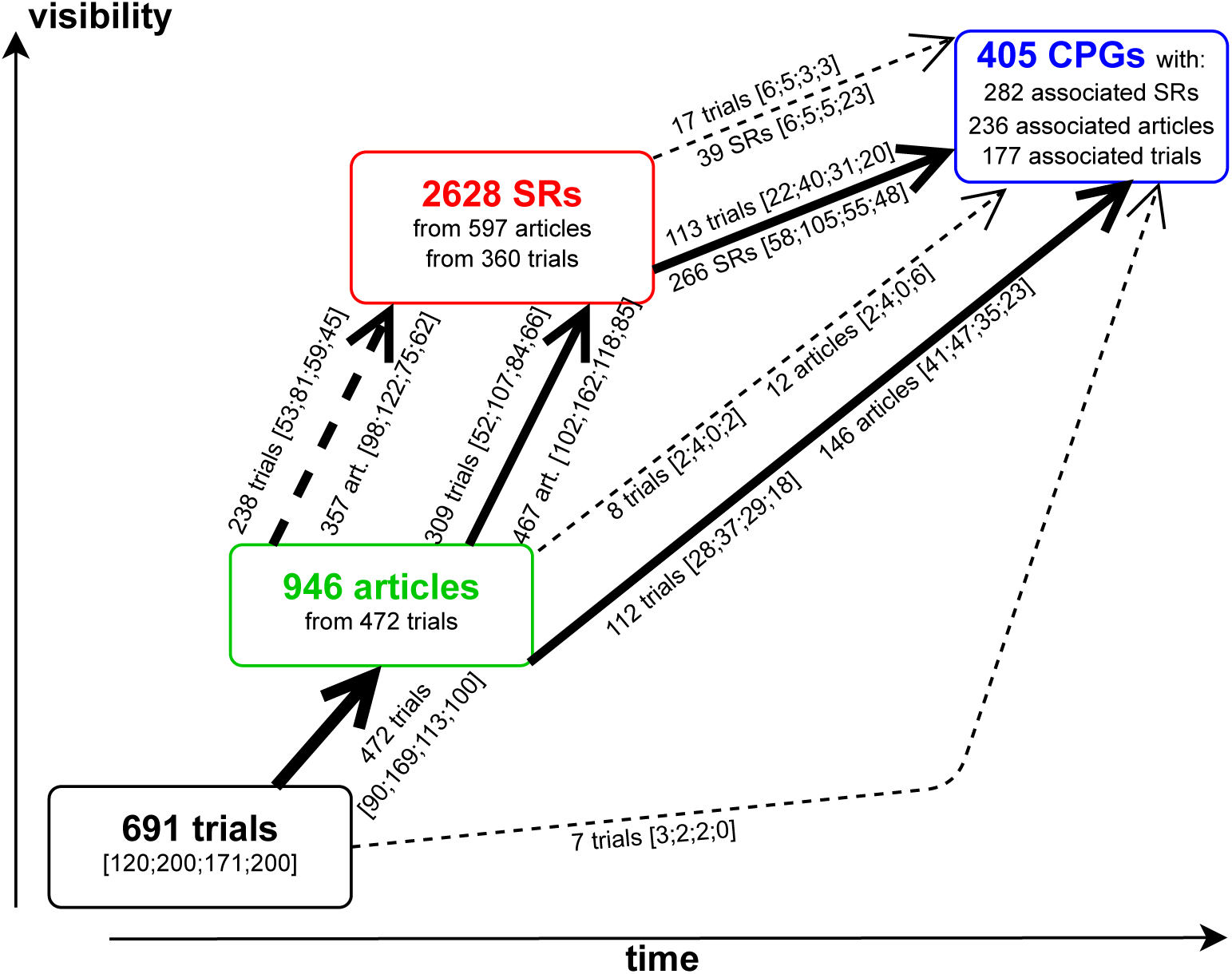
Flowchart data representation. Total number of trials, published articles, SR citing these articles and CPGs citing the articles or the SRs. Dotted arrows indicate non-inclusion in the citing entity, solid arrows indicate inclusion. Paths which were taken by a considerable (>100) number of trials are thicker. Since for example trials with articles can be cited with their articles in SRs and be excluded as well as included, the sum of 238 trials and 309 trials merely represents an upper bound for the total number of distinct trials being associated with SRs (360). The same type of argument applies to the other paths. Based on the concept for assessment of research impact as described by [3, 4].

### Impact-on-CPGs and impacted-CPGs per trial

Fitting the multivariable negative binomial regression model showed that the impact on CPGs per trial was similar across the cohorts, with the exception of the international ISTs (Table 1). Those underperformed in comparison to the German IITs with an incidence rate ratio of 0.42 (95% CI [0.19, 0.87], p-value 0.023), while correcting for the covariates drug trial, number of primary outcomes above median and study size above median. Also, a study size above the median of 150 was associated with a significantly higher impact-on-CPGs (IRR 5.13 CI [2.90, 9.20], p-value <0.001). The effect of study size remained when modeling the outcome impacted-CPGs per trial with an IRR of 3.10 (CI [2.05, 4.71], p-value <0.001). The effect of international ISTs was not significantly inferior for this outcome. As sensitivity analysis we also modeled associated-CPGs per trial with the exact same method and covariates as shown in Table 1 yielding very similar results: significant inferiority of international ISTs (IRR 0.55, CI [0.31, 0.94], p-value 0.033) and significant superiority for trials with study size above median (IRR 3.02 CI [2.06, 4.47], p-value <0.001). An additional analysis with the same covariates, but without dichotomizing number of primary outcomes and study size yielded similar results, with significant effects for the same explaining variables with similar directions.

**Tab. 1:** Multivariable negative binomial regression models for a) Impact-on-CPGs and b) Impacted-CPGs per trial. The observed time per trial was taken into account as an offset term after log-transformation.

|  | Impact-on-CPGs per trial |  |  | Impacted-CPGs per trial |  |  |
| --- | --- | --- | --- | --- | --- | --- |
| Covariates | IRR | 95% CI | p-value | IRR | 95% CI | p-value |
| **Cohort** |  |  |  |  |  |  |
| German IITs | — | — |  | — | — |  |
| International IITs | 1.31 | 0.59, 2.82 | 0.5 | 1.36 | 0.76, 2.42 | 0.3 |
| German ISTs | 1.22 | 0.53, 2.78 | 0.6 | 1.30 | 0.71, 2.39 | 0.4 |
| International ISTs | 0.42 | 0.19, 0.87 | 0.023 | 0.61 | 0.34, 1.08 | 0.10 |
| **Drug trial** |  |  |  |  |  |  |
| no | — | — |  | — | — |  |
| yes | 0.81 | 0.48, 1.36 | 0.4 | 1.08 | 0.73, 1.58 | 0.7 |
| **Study size** |  |  |  |  |  |  |
| <= 150 | — | — |  | — | — |  |
| > 150 | 5.13 | 2.90, 9.20 | <0.001 | 3.10 | 2.05, 4.71 | <0.001 |
| **Number of primary outcomes** |  |  |  |  |  |  |
| > 1 | — | — |  | — | — |  |
| 1 | 0.53 | 0.29, 0.92 | 0.031 | 0.65 | 0.41, 1.00 | 0.059 |

IRR = Incidence Rate Ratio, CI = Confidence Interval

### Time-to-guideline impact

For the analysis of the time-to-guideline impact we illustrate Kaplan-Meier curves for the cumulative event probability over time for the four groups in Figure 5. There is a statistically significant difference between the groups (*p* = 0.039), indicating that the probability of events occurring varies across them. Notably, international ISTs appear to have a lower cumulative event probability compared to the other three groups.

**Fig. 5:**
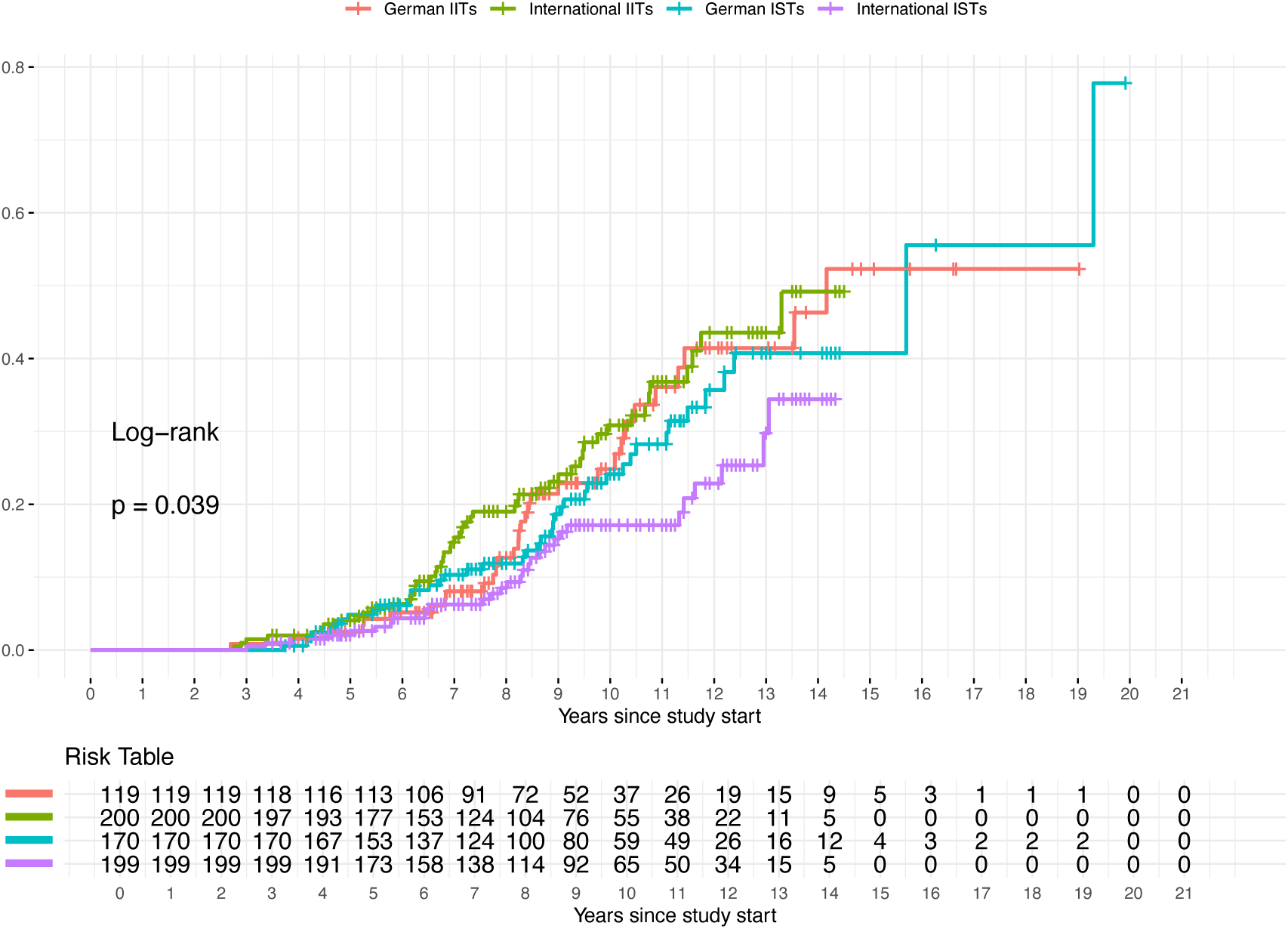
Kaplan-Meier estimates for the probability of having a guideline impact are displayed as the cumulative event. The time-to-event was the time from trial start to the publication of the first CPG impacted by a trial. If a trial did not impact a CPG until the end of the observation period, it was treated as right censored with the censoring date 31st August 2019. An event happened for 153 of 691 studies. The risk table shows the trials still at risk at each year since study start stratified by cohort.

To take the effect of covariates into account we also fitted a Cox proportional hazards model for the time-to-guideline impact (Table 2). Again, the international ISTs seem inferior with a hazard ratio (HR) of 0.61 (95% CI [0.37, 1.00]), though with a p-value of 0.052. Trials bigger than the median study size seemed superior with an HR 2.52 (95% CI [1.77, 3.59], p-value <0.001). Repeating the analysis without dichotomizing study size and the number of primary outcomes produced similar results. Visual inspection of the Schönfeld residuals for each explaining variable in the Cox model revealed no trends over time thus not giving indication that the proportional hazard assumption might be violated [6].

**Tab. 2:** Multivariable Cox proportional hazards model for the time from trial start to guideline impact. For definition of event and time-to-event see also the caption of Figure 5.

| **Covariates** | **HR** | **95% CI** | **p-value** |
| --- | --- | --- | --- |
| **Cohort** |  |  |  |
| German IITs | — | — |  |
| International IITs | 1.35 | 0.86, 2.12 | 0.2 |
| German ISTs | 1.03 | 0.64, 1.65 | >0.9 |
| International ISTs | 0.61 | 0.37, 1.00 | 0.052 |
| **Drug trial** |  |  |  |
| No | — | — |  |
| Yes | 1.02 | 0.73, 1.42 | >0.9 |
| **Study size** |  |  |  |
| <= 150 | — | — |  |
| > 150 | 2.52 | 1.77, 3.59 | <0.001 |
| **Number of primary outcomes** |  |  |  |
| > 1 | — | — |  |
| 1 | 0.86 | 0.58, 1.25 | 0.4 |

*HR = Hazard Ratio, CI = Confidence Interval*

## Discussion

The goal of this work was to comprehensively assess the impact of RCTs on CPGs. We provided a framework for assessing visibility and relevance of RCTs by defining several measures for impact. Based on these measures, we estimated the number of impactful guideline citations and the time-to-guideline impact from the IMPACT data set. Our findings indicate strong evidence that larger trials have more impact on CPGs and impact CPGs faster, while international ISTs seem to have less impact and are slower to do so. It remains unclear, however, why the international ISTs underperform the other cohorts in our study. Had there been a significant performance gap between German and international RCTs, we would also expect to see a difference between German and international IITs. With analogous reasoning it is unlikely that there is a large performance gap between IITs and ISTs. As for potential confounders we did account for several trial baseline characteristics in our analysis: study size, number of primary outcomes, drug trial, study phase and medical domain. In the investigation of the publication process, Blümle et al. [5] found the international IITs, the German ISTs and the international ISTs to be on par with the German IITs in terms of publication rate and even to be superior for the time-to-publication. This makes it unlikely for either of these factors to explain the effects we found in this study. If only journal articles were considered as a relevant form of publication, the international ISTs would only be at a slight disadvantage with rates of 68% [75%, 84%, 66%, 51%] respectively. It could be argued that this data carries a potential risk of domestic preference, as the search included both English-language guidelines and specifically German-language guidelines. This approach might favor German RCTs or, at the very least, make it less likely for guideline authors to be unaware of an RCT conducted within their own country and field of expertise. Such a preference could potentially obscure any inferiority in the German cohorts. To address this, we reran the analysis, excluding all German-language CPGs. This analysis did not indicate any inferiority in any outcome for any of the German cohorts. The proposed method to assess impact on CPGs is objective and easy to implement. One might wish for an even more precise method, for example by only considering citations in guideline recommendations, by weighting citations by the strength of recommendations or the level of evidence assigned to the cited reference or by weighting the citations by the guideline-quality. While we considered all of these, there are compelling reasons not to do so. Regarding the choice not to use the citations in recommendations only: this approach is not feasible in reality, because for it to work, the recommendations would have to be clearly labelled as such in the guidelines and the underlying evidence would somehow have to be mentioned directly with the recommendations. Only 29% of the guidelines in our data set did this unambiguously. A similar small portion of guidelines provides strength of recommendations or level of evidence, making this approach impractical as well. Lastly, while tempting in theory, assessing guideline quality is also impractical. While to the best of our knowledge two instruments exist which are sufficiently validated for research purposes (AGREE II [7] and DELBI [8]), they are unsuited for comparing guidelines, time-consuming and demand different researchers to conduct the evaluation independently [9]. For future studies assessing the impact on CPGs it might be sufficient to simply measure the number of associated CPGs per trial. This would reduce the resources needed to conduct the study and yielded comparable results for our set of RCTs. However, as Figure 4 suggests, it seems worthwhile to distinguish between exclusion and inclusion in SRs on the basis of the scale alone. Also, the amount of time needed by the investigator does not change drastically between identifying the guideline, confirming citation of the cited references and extracting the publication-date vs. additionally searching for all the citations of the cited reference and adding those up. Since there is a wide variety in how an RCT can be important for clinical practice, we did not exclude paths with method articles and did not outright exclude direct paths from RCTs to guidelines when defining the path-attribute *impactful*. One example of this could be that an RCT needed to be changed due to unforeseen treatment side effects in a subcohort. This would be valuable information for the practitioner evaluating different treatment choices. Regarding our choice to investigate the time-to-guideline impact we also considered to investigate the time from trial completion to guideline-impact. This however proved to be difficult due to problems with the completion date. The completion date was extracted from the registries, but the registries did not have common definitions for the completion date, and it was missing in 11 cases. To name just one example where the notion of time from completion to guideline impact is difficult and ambiguous: one trial in our data was a large drug trial evaluating the effect of a biological on psoriasis. It consisted of four different stages: placebo-controlled phase, placebo crossover + active treatment phase, randomized withdrawal phase and five years of safety observation. It reported trial completion after the safety observation, but already published two highly cited papers before that and did also reach guideline-impact 1.5 years before the end of the safety observation period. Another initially attractive idea might be to investigate the time from article to guideline-impact. However, this would be equivalent to studying a subcohort with a high risk of selection and survivorship bias, as only a select portion of RCTs published an article. One problem of this study might be that it could be underpowered. For the initial IMPACT study a power analysis was conducted with the time-to-publication analysis in mind and the sample sizes were chosen to achieve 90% power for a HR < 0.625 or HR > 1.6 assuming a long-term event rate of 50%. The observed long term event rate for the event guideline-impact was only 23%. Another hint for a problem with our time-to-guideline analysis might be that the Kaplan-Meier estimates did not form a plateau whilst there were still a relevant number of RCTs at risk for the event. We chose to model the impact on the CPGs and the impacted CPGs using negative binomial models rather than Poisson models, because of the overdispersion in the data. The data generation process gave us no reason to use zero-inflated models, whereas non-zero-inflated negative binomial models, as used in this study, has been shown to be able to handle a large number of zeros quite well [10, 11]. This study is limited by the technical inability to conduct a search for SRs citing the trials directly and not only their articles. This was because SRs were identified via the “cited by” functionality of PubMed and Web of Science. With this method, it was only possible to search for SRs citing known DOIs. This might be a disadvantage for the commercial cohorts as these did publish their results in the registry only more often [2%;4%;20%;32%][5]. In addition, there might be important confounders like other RCT baseline characteristics that we did not account for. For future research, it seems recommendable from our data to control for country of primary study site, sponsor type, study size and, possibly, study phase. Another limitation could be that German IITs might be subject to selection bias and the results might not be widely extendable. This cohort is a selection of RCTs that (i) got approval in Germany and (ii) were able to gain funding. Assuming that these trials are of a particular quality and that their results are of particular interest, a strong influence is less surprising. However, regarding the first argument, these are multi-center studies that also had to gain approval in other countries. Furthermore, in a 2009 survey of pharmaceutical companies, Ruppert and Pfeiffer [12] essentially conclude that the approval process in Germany, at least in Europe in the 2000s, represented a positive factor for choosing Germany to conduct a RCT. For the initiation of IITs in Germany the bureaucratic hurdles of the 2000s seem to have posed a considerable problem [13], however, the question remains whether this possible discrepancy between IITs and ISTs was specific to Germany. Speaking to the other factor of selection – funding by DFG or BMBF – this would not impact the comparison between the German ISTs and international ISTs. Ergo one would still be able to conclude that at least in the case of ISTs the German RCTs do not perform inferior in terms of impact on CPGs.

## Conclusion

German RCTs seem to perform at least comparable in guideline impact as well as time-to-guideline impact in comparison to international RCTs. It is unsatisfactory that only 29% of the guidelines make a clear link between their recommendations and the underlying evidence. On one hand, this hinders conducting detailed meta-studies like this one, but more importantly it makes it challenging for clinicians to form informed decisions and rapidly identify the most relevant evidence to their problem. It should become a minimum standard for a CPG to cite explicitly and directly in key places. Due to their unstandardized form, CPGs are often hard to find especially for foreign stakeholders or outright impossible to access from foreign countries. Furthermore, it is often not clear whether they are peer-reviewed or not. It should become standard practice for CPGs to be published as an article in a peer-reviewed journal, possibly in several languages. This would also make them easier to locate, as those articles would be listed in biomedical literature databases, such as Medline. Given that a considerable number of ISTs only publish their results in registries, it should become standard practice for SRs and CPGs to also search for relevant evidence in primary trial registries. Overall, the fact that only 25% of trials in our data are associated with a CPG remains unsatisfactory. It is highly unlikely that there are no CPGs for the other RCTs for which their findings would have been relevant. Even though this might in part be explained by only 68% of trials being published as a journal article, there are still 42% of all trials which are published as an article but were not associated with a CPG. This hints to a need for improvement in RCT visibility.

## Data Availability

The datasets used and/or analyzed during the current study are available from the corresponding author on reasonable request.

## Abbreviations

AWMF: Arbeitsgemeinschaft der Wissenschaftlichen Medizinischen Fachgesellschaften (Association of the Scientific Medical Societies). https://www.awmf.org/leitlinien/leitlinien-suche.html
BMBF: Bundesministerium für Bildung und Forschung (Federal Ministry of Education and Research). https://www.bmbf.de
CI: Confidence Interval
DFG: Deutsche Forschungs-gemeinschaft (German Research Foundation) https://www.dfg.de/
DOI: Digital Object Identifier
DRKS: Deutsches Register Klinischer Studien (German Clinical Trials Register). https://www.drks.de
EUCTR: EU Clinical Trials Register. https://www.clini-caltrialsregister.eu/
Google scholar: Web search engine providing scholarly literature. https://scholar.google.com
HR: Hazard Ratio
IITs: Investigator Initiated Trials
IRR: Incidence Rate Ratio
ISRCTN registry: International Standard Randomized Controlled Trials Number registry. http://www.isrctn.com
ISTs: Industry-Sponsored Trials
LIVIVO: Interdisciplinary search engine for literature and information in the field of life sciences, run by ZB MED – Information Centre for Life Sciences. https://www.livivo.de
Medline: Medical Literature Analysis and Retrieval System Online. Bibliographic database of life sciences and biomedical information. Accessible e.g. via the search engine PubMed
NICE: National Institute for Health and Care Excellence. https://www.nice.org.uk/guidance/
PubMed: Free search engine accessing primarily the MEDLINE database. https://www.ncbi.nlm.nih.gov/pubmed/
RCT(s): Randomized controlled trial(s)
SR(s): Systematic review(s)
TRIP: Turning Research Into Practice. https://www.tripdatabase.com/
Web of Science: formerly https://apps.webofknowledge.com, now https://www.webofscience.com/wos/woscc/basic-search

## Supplementary Information

Additional file 1: Transition plots of three additional exemplary trials highlighting the diversity within our dataset

## Declarations

### Ethics approval and consent to participate

Not applicable.

### Consent for publication

Not applicable.

### Authors’ contributions

AB and MS designed the IMPACT study. MH, NB and MS designed this study. NB provided comprehensive guidance throughout the study. MH searched for the guidelines and extracted the data. MH analyzed the data and conducted the statistical analyses. MH wrote the manuscript. NB and AB substantially revised the manuscript. All authors read and approved the final version of the manuscript before submission.

## Acknowledgements

We would like to thank everyone who worked on the IMPACT study.

