## Supplementary Material for "Investigator-initiated versus industry-sponsored trials – Visibility and relevance of randomized controlled trials in clinical practice guidelines (IMPACT)"

### Additional file

In this supplement the lifecycle of three more trials is displayed to demonstrate the observed diversity in our data.

1. Transition plot of a trial with no impact on CPG despite several publications and citations in SRs

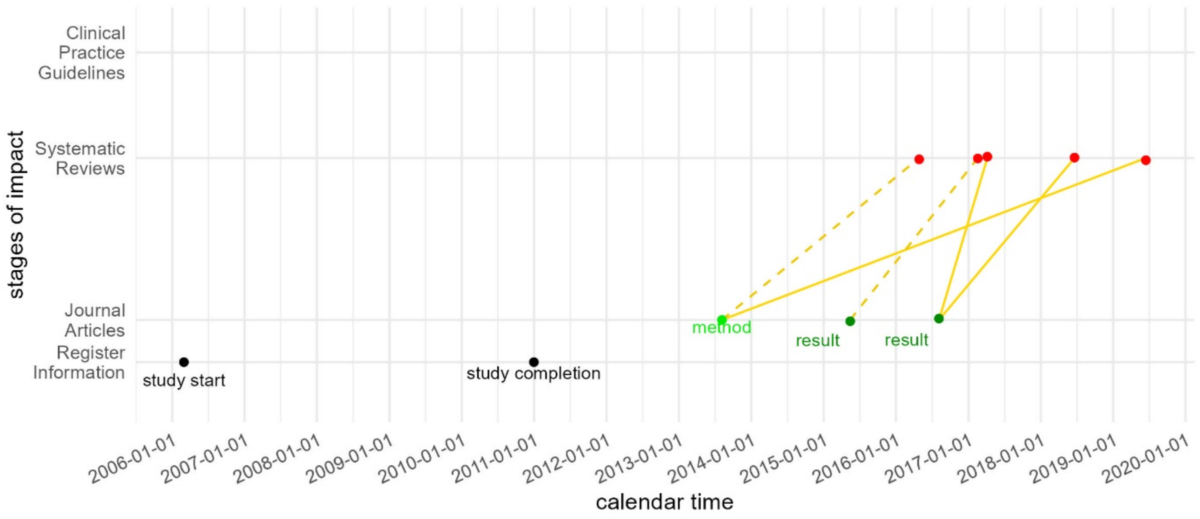


The above trial was completed and published three papers. These were cited and included in SRs (first in 2016 and 2017 respectively). Still, no CPG cited any of these entities, thus this trial had no impact on CPGs.

2. Transition plot of a trial with CPG impact despite publishing only one article

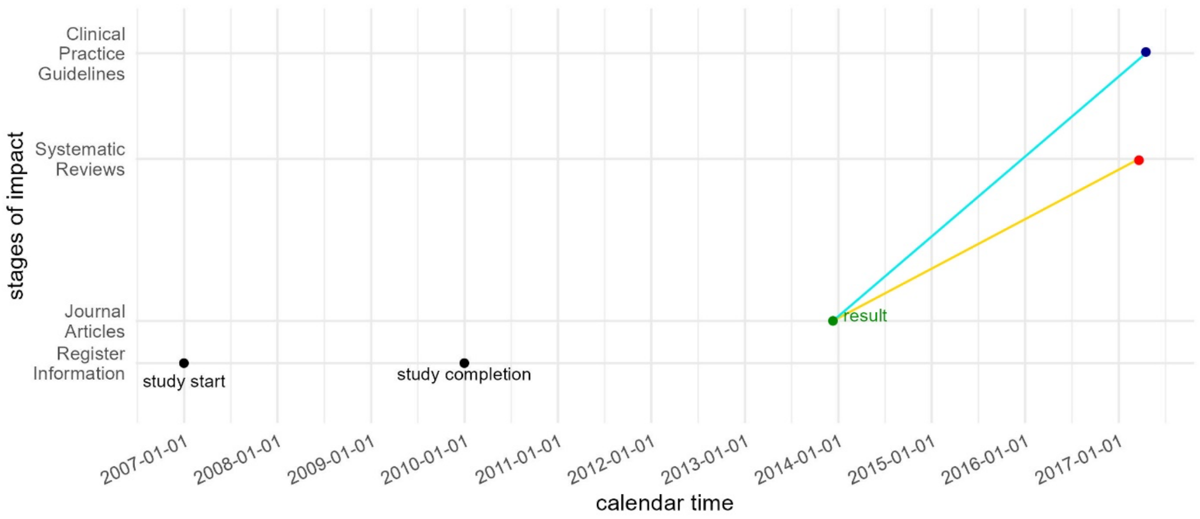


This trial gained impact in CPGs even though it only published one article. The trial was also included in an SR. Hence it is an example how one article might be sufficient for impact on CPGs.

3. Transition plot of a trial with several citations, incl. a CPG, yet no guideline impact

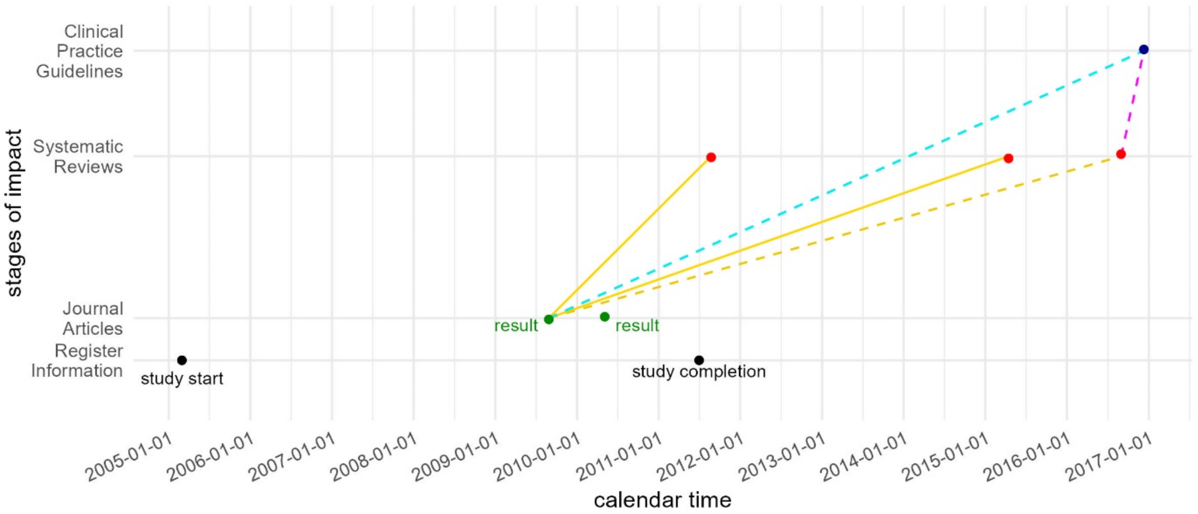


The last example features a trial, which was included in two SRs and was cited directly and indirectly in a CPG, but still gained no impact on CPGs, because the citation in CPG was not impactful. This trial also exhibits one common feature in our data, that the first result article seems to commonly be the most cited article of a trial. This trial highlights the nuance in our approach to define guideline impact.
